# Interplay between Sex and Disease Burden in Huntington’s Disease: Clinical and Neuroimaging Perspectives

**DOI:** 10.1101/2024.10.31.24315900

**Authors:** Jingwen Yao, Grayson Feng B.S., Guowen Shao M.S., Adys Mendizabal, Donatello Telesca, Kaiying Fang, Lola Ibragimova, Juwairia Shoaib, Melanie A. Morrison, Janine M. Lupo

## Abstract

**Background:** Huntington’s disease (HD) is a progressive neurodegenerative disorder caused by a cytosine-adenine-guanine (CAG) repeat expansion in the hungtintin gene. The disease exhibits sex-related differences in symptomatology and disease progression, but the effect on brain structural biomarkers and the interaction between sex and disease burden remain underexplored.

**Objectives:** To investigate the interplay between sex and disease burden on clinical measures and neuroimaging biomarkers in HD.

**Methods:** We retrospectively analyzed data from Enroll-HD, TRACK-HD/ON, PREDICT-HD, and IMAGE-HD studies, including a combined dataset of 19,738 participants with CAG>=40. Linear mixed models were employed to evaluate the influence of sex and the sex-disease burden interaction on clinical evaluations (Unified Huntington’s Disease Rating Scale and Problem Behaviors Assessment) and neuroimaging biomarkers (striatal volumes and cortical thickness), with CAG-Age Product Score (CAPS) used as a proxy for disease burden, while controlling for covariates.

**Results:** Female participants exhibited less pronounced striatal atrophy and cortical thinning with increasing CAPS (caudate: *β_male/female_*=–3.462/–2.935, p<0.05; putamen: *β_male/female_*=–4.775/–3.908, p<0.01). Regarding clinical measures, females experience greater motor decline with increasing CAPS (Total Motor Score: *β_male/female_*=3.024/3.244, p<0.0001), greater cognitive decline (Symbol Digit Modalities Test : *β_male/female_*=–34.89/–38.91, p<0.0001), and greater functional decline (Total Functional Capacity: *β_male/female_*=–6.449/–6.817, p<0.05; Independence Scale: *β_male/female_*=–32.44/– 34.99, p<0.001).

**Conclusions:** The sex-CAPS interaction significantly impacts both clinical and neuroimaging biomarkers of HD, underscoring the importance of incorporating sex-specific considerations into the clinical staging and management of HD.

## INTRODUCTION

Huntington’s disease (HD) is a progressive neurodegenerative disease caused by a cytosine-adenine-guanine (CAG) repeat expansion in the hungtintin (HTT) gene on chromosome 4. The number of CAG repeats is closely linked to symptom onset and disease severity ^1^. The mutated protein accumulates over time, causing the gradual emergence of involuntary movements and other motor disorders including chorea, dystonia, and dysarthria, a decline in cognitive function including executive dysfunction, and neuropsychiatric issues such as depression and suicidal ideation. A higher number of repeats typically result in earlier symptom onset and faster disease progression ^2^.

With an autosomal dominant pattern of inheritance, research on sex differences in HD has been limited. Nonetheless, recent evidence has revealed sex-related disparities in disease prevalence, symptomatology, and progression rate, suggesting sex plays a critical role in HD phenotype and disease trajectory^3–8^, with implications for differential disease prognostication and patient management. Analyses of data from the REGISTRY and ENROLL-HD databases indicated that female HD subjects exhibit more severe motor and functional impairments, worse depression symptoms, and faster rate of progression rates ^4–6^. Additionally, motor symptoms primarily contribute to functional disability in females, while cognitive deficits were more explanatory of functional variation in males ^4^. Lastly, a national epidemiological study involving 67 million US residents identified a modest but significant difference in HD prevalence between women and men (7.05 vs. 6.91 per 100,000) ^9^.

Investigations into sex differences in HD biomarkers beyond clinical measures are scarce. One study examined the impact of cerebrospinal fluid neurofilament light chain (NfL) levels on clinical outcomes and brain atrophy in HD, identifying a significant interaction between NfL levels and sex. Higher NfL concentrations correlated with more pronounced atrophy and clinical severity in females ^8^. This finding mirrors the observations of sex-differential associations between biomarkers or genetic risk factors and disease progression in other neurodegenerative disorders. For instance, the ε4 allele of the apolipoprotein E gene (APOE4), a significant risk factor for sporadic and late onset familial Alzheimer’s disease, showed a stronger correlation with disease onset, memory decline, and cognitive impairment in females compared to males [8, 9].

While previous studies have highlighted sex differences in HD manifestation independent of disease burden, the potential of sex as an effect modifier in HD has yet to be evaluated in large datasets. The development of HD Integrated Staging System (HD-ISS) ^12^ represents a significant advancement in standardizing disease staging to facilitate clinical research and interventional studies. Importantly, neuroimaging markers (caudate and putamen volumes) are incorporated as landmarks of Stage 1 HD to capture the transition into neurodegeneration early in the disease course. This raises the question of whether brain atrophy measures will reveal similar patterns of sex differences and potential interactions with disease burden. Additionally, Total Motor Score (TMS) and Symbol Digit Modalities Test (SDMT) have been selected as landmarks of Stage 2, while Total Functional Capacity (TFC) and Independence Scale are used as landmarks of Stage 3, the final stage of the disease.

The interaction between sex and disease burden on these clinical and biological markers in HD could have profound implications for patient staging within the HD-ISS framework. To improve our understanding of these interactions and assess the necessity of sex-specific considerations in the HD-ISS and clinical monitoring of disease progression, this study aims to investigate the interplay between sex and disease burden on clinical measures and neuroimaging biomarkers, by analyzing a comprehensive dataset compiled from four HD studies.

## MATERIALS AND METHODS

### Datasets and study population

We retrospectively analyzed data from four observational HD study datasets: Enroll-HD^13^, TRACK-HD/ON^14–16^, PREDICT-HD^17^, and IMAGE-HD^18,19^. Specifically, we included data from Enroll-HD periodic dataset 6, extracted on November 4, 2022, containing data from 95,040 visits of 25,550 participants. PREDICT-HD dataset includes 7700 visits from 1485 participants from 2001 to 2017. TRACK-HD/ON dataset comprises 2110 visits from 446 participants, and the IMAGE-HD dataset includes 3-year follow-up data from 108 participants. Additional information on these datasets is included in **Supplementary Table 1**. All data were obtained and maintained with local ethical approval. Data from all four datasets were combined to generate a final comprehensive dataset with unified parameter coding for analysis.

Different HD studies used varying criteria for HD group assignment. We unified the participant subgroup definition as follows. Participants with CAG repeat length >=40 were included in HD groups, aligned with the latest HD staging recommendation ^12^. HD subjects were categorized as manifest with a TMS > 10 and/or a diagnostic confidence level = 4 at each visit. Otherwise, they were determined as premanifest (premotor) HD. Demographic information, including sex, age, race/ethnicity, education level, and substance use, were collected to serve as covariates in the analysis, as these variables have been associated with differences in clinical measures in HD ^20^. Race/ethnicity groups included Caucasian, Black/African American, Hispanic/Latino, and Others. Education levels were determined based on the International Standard Classification of Education. Participants with any substance use or substance use history, including alcohol, tobacco, or drug, were classified as with substance use. The CAG-Age Product score (CAPS) was calculated as a proxy of HD disease burden, using the equation: CAPS = Age×(CAG-33.66)/ 432.3326 ^21^.

We compared the CAG repeat length, baseline CAPS, and demographic factors between male and female participants. For continuous variables, a one-sample Kolmogorov-Smirnov test was first performed to examine the normality of data distribution, with p < 0.05 indicating non-normal distribution. Normally distributed samples were compared using a Student’s t-test; otherwise, a Mann-Whitney U-test was used. Categorical variables were compared using a chi-square test.

### Clinical assessment

We included the following motor, functional, neuropsychiatric, and cognitive assessments in the analysis: (1) Motor: Unified Huntington’s Disease Rating Scale (UHDRS) TMS; (2) functional: UHDRS TFC, UHDRS Independence Scale; (3) neuropsychiatric: Problem Behaviors Assessment – Short (PBA-S) depression, irritability, psychosis, apathy, and executive function scores; (4) cognitive: SDMT. Among the metrics, TMS and SDMT were available in all datasets. TFC and Independence Scale were available in Enroll-HD, TRACK-HD/ON, and PREDICT-HD studies. PBA-S scores were only available in Enroll-HD and PREDICT-HD datasets.

### Imaging analysis

We evaluated neuroimaging-based volumetric biomarkers using T_1_-weighted structural MR images in the PREDICT-HD, TRACK-HD/ON, and IMAGE-HD datasets (3835 scans from 1303 HD participants). All three studies utilized a three-dimensional T_1_-weighted magnetization-prepared rapid acquisition with gradient echo (MPRAGE) or volumetric spoiled gradient echo acquisition (SPGR) at 1.5T or 3T. Representative imaging parameters documented by the studies are included in **Supplementary Table 2**. To ensure consistency in processing, all imaging data were processed with FreeSurfer (version 6.0.0) to extract caudate, putamen, and estimated intracranial volume (eTIV), and perform cortical thickness evaluations (**Supplementary Figure 1**). Due to the high number of images, we adopted an automated quality assurance strategy, omitting imaging data with a Euler number lower than 25th percentile – 1.5× the interquartile range of the sample. Euler number provided by the Freesurfer toolbox summarizes the topological complexity of the reconstructed surfaces, as calculated by the sum of the vertices and faces subtracted by the number of edges, where a topologically perfect reconstruction should result in a surface with Euler number of 2 ^22^, with smaller numbers representing greater level of segmentation defects. This automated quality control approach has shown high correlation with human rater visual quality control and can identify unusable images by human raters with area under the curve of 0.98-0.99 ^23^.

### Statistical modeling of sex-disease burden interaction on HD markers

To understand the influence of sex and the sex-disease burden interaction on HD markers, we employed two linear mixed model to fit the longitudinal data, with the second model containing an additional interaction term between sex and CAPS compared to the first model.

Model 1: *y_im_* = *β*_0_ + *β*_1_*Age_i_* + *β*_2_*Substance_i_* + *β*_3_*Education_i_* + *β*_4_*Race_i_* + *β*_5_*Sex_i_* + *β*_6_*CAPS_i_* + *β*_7_*Days_i_* + *β*_8_(*Days_i_* * *Sex_i_*) + *β*_9_(*Days_i_* * *CAPS_i_*) + *b*_0*m*_ + *b*_1*m*_*Days_im_* + *∈_im_;*

Model 2: *y_im_* = *β*_0_ + *β*_1_*Age_i_* + *β*_2_*Substance_i_* + *β*_3_*Education_i_* + *β*_4_*Race_i_ + β*_5_*Sex_i_* + *β*_6_*CAPS_i_* + *β*_7_*Days_i_* + *β*_8_(*Days_i_* * *Sex_i_*) + *β*_9_(*Days_i_* * *CAPS_i_*) + *β*_10_(*Sex_i_* * *CAPS_i_*) + *b*_0*m*_ *+ b*_1*m*_*Days_im_* + *∈_im_,*

where i = 1, 2, …, N_visits_, and m = 1, 2, …, N_participants_. *β* represents the fixed effect coefficients, and *b*_O*m*_ and *b*_1*m*_ represent the random effects of intercept and slope with respect to days from baseline for each subject *m. y* represents the dependent variable, either clinical assessment score or imaging biomarker. Fixed effect variables included subject age, substance use, education level, race, sex, CAPS, days from baseline, and the interaction between sex and time interval, CAPS and time interval, as well as between sex and CAPS in the second model. For imaging biomarkers, additional fixed effect variables of MRI field strength and eTIV were included to account for acquisition and head size differences. Sex, race, substance use, and MRI field strength were treated as categorical variables. To decouple the effect of time from baseline and subject’s age, baseline age and CAPS were used for all time points within each subject. TMS scores were log-transformed using log (*TMS* + 1) to reduce skewness before model fitting. Missing data in substance use and education level (< 1%) were imputed with the median value from the entire population.

To evaluate whether the addition of sex-CAPS interaction improves model fitting, we conducted a likelihood ratio test comparing Model 2 against Model 1. If Model 2 resulted in a better fitting compared to Model 1 with the likelihood ratio test *p*-value < 0.05, Model 2 was used to obtain the statistics, otherwise, Model 1 was used. The final model fitting statistics were adjusted using the Benjamini-Hochberg (BH) procedure to control the false discovery rate. A *p*-value of < 0.05 was considered significant.

### Cortical thickness evaluation

Cortical thickness evaluation was performed using FreeSurfer on the T_1_-weighted images. The processed brain surfaces were smoothed using a 10 mm full-width half-maximum filter and aligned to a standard space. To identify significant differences in cortical thickness, the previously described linear mixed (Model 2 with MRI field strength as an additional fixed variable) was applied at the vertex level. The threshold for statistical significance was set at *p* < 0.05, with adjustments for multiple comparisons performed using Monte Carlo permutations.

### Impact of sex on HD-ISS staging

To evaluate whether the probability of being assigned different disease stages varies between male and female HD subjects, we calculated the percentage of male and female subjects assigned to each stage within specific age ranges and CAG repeat lengths. Age was stratified using 5-year sliding windows, and CAG repeat length was stratified at each individual repeat length. The assignment of disease stage was based on the operational age-based cut-off biomarker values as described in the HD-ISS ^12^. These staging proportions were then compared using a chi-square test. The BH procedure was applied to control the false discovery rate across age and CAG ranges. Due to the limited number of subjects with imaging biomarkers after stratifying for age and CAG repeat length, we evaluated only Stage 2 and Stage 3 assignments based on the stage-specific landmarks and did not assess the stage as the sum of binary condition variables, as described by the original method ^12^, where stage assignment can be scored as the sum of the binary indicators of each stage condition.

### Data availability statement

Data used in this work were generously provided by the participants in the Enroll-HD, PREDICT-HD, TRACK-HD/ON, and IMAGE-HD studies and made available by CHDI Foundation, Inc.

## RESULTS

### Population Demographics

Participant demographics are detailed in **Table 1**. The cohort comprises 19,738 individuals, including 8,975 males and 10,763 females. The subset of data with imaging included 3640 scans from 1262 subjects after quality assurance (195 scans excluded). Significant differences in age, substance use, and race/ethnicity distributions between males and females were observed, which we included as covariates in our mixed model. Education level, though not significantly different between sexes, was included due to its impact on SDMT ^24^ and TFC ^20,25^.

**Table 1.** Participant demographics.

| Demographic variables |  | All Subjects | Male | Female | *p-value* |
| --- | --- | --- | --- | --- | --- |
| Age (mean ± SD) | | 47.4 ± 13.8 | 48.2 ± 13.6 | 46.8 ± 13.9 | **** |
| CAG (mean ± SD) | | 43.7 ± 3.5 | 43.7 ± 3.5 | 43.7 ± 3.5 | n.s. |
| CAPS (mean ± SD) | | 1.0 ± 0.3 | 1.1 ± 0.3 | 1.0 ± 0.3 | **** |
| Education Level (mean ± SD) | | 3.7 ± 1.2 | 3.6 ± 1.2 | 3.7 ± 1.2 | n.s. |
| Substance Use (%) |  | 71.2% | 76.9% | 66.6% | **** |
| HD group (%) | Manifest | 65.0% | 68.8% | 61.7% | **** |
|  | Premanifest | 35.0% | 31.2% | 38.3% |  |
| Race / ethnicity Groups (%) | Caucasian | 93.1% | 93.6% | 92.6% | ** |
|  | African American / Black | 0.6% | 0.6% | 0.6% |  |
|  | Hispanic / Latino | 2.7% | 2.2% | 3.1% |  |
|  | Other | 3.6% | 3.5% | 3.7% |  |
| Number of Imaging Sessions (N) |  | 3640 | 1442 | 2198 | / |
| MRI Field Strength (%) | 1.5T | 38.4% | 36.3% | 39.7% | / |
|  | 3T | 61.6% | 63.7% | 60.3% |  |

Abbreviations: SD: standard deviation; CAG: cytosine-adenine-guanine; CAPS: CAG repeat length-Age
Product Score; HD: Huntington's disease; \*\*\*\*: $p$ -value $< 0.0001$ ; \*\*: $p$ -value $< 0.01$ ; n.s.: not significant.

### HD-ISS Stage 1 landmarks – caudate and putamen volumes

Volumetric analyses revealed significant associations between sex and subcortical structure volumes, with females exhibiting smaller putamen (p < 0.001) volumes after controlling for covariates. The sex-CAPS interaction significantly influenced both caudate (p < 0.05) and putamen (p < 0.01) volumes, suggesting less CAPS-mediated atrophy in females (**Figure 1A**). In manifest HD subjects, the sex-CAPS interaction remained significant for caudate volume only (p < 0.05), while premanifest subjects showed significant sex differences in putamen volume (p < 0.05).

**Figure 1.**
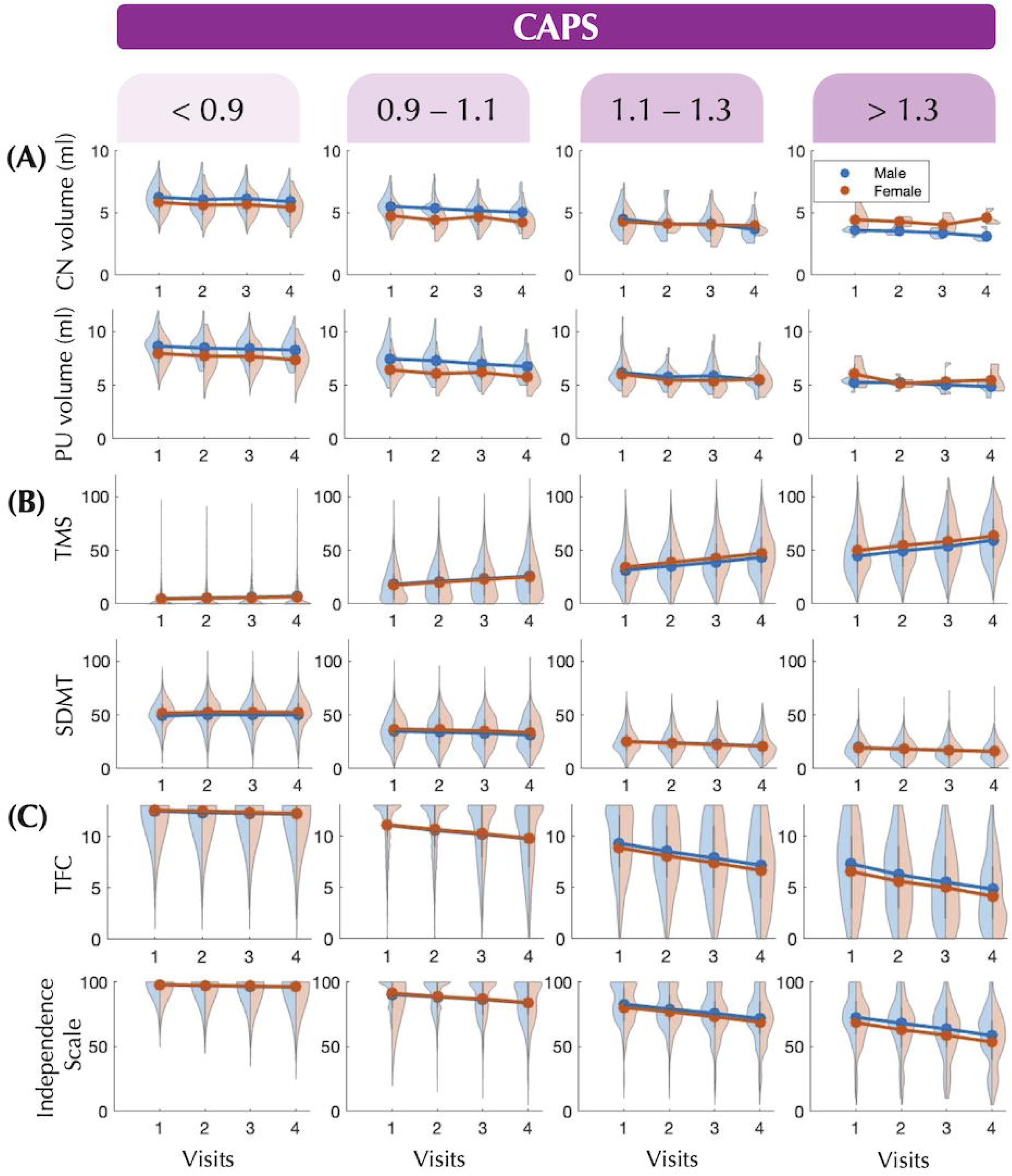
Longitudinal changes in HD-ISS landmarks compared between male and female HD gene carriers over visit 1-4. Violin plots of HD-ISS Stage 1-3 landmarks (A-C) are illustrated, with line plots showing the mean landmark values within the timepoints.

### HD-ISS Stage 2 landmarks – TMS and SDMT

Significant sex-disease burden interactions were observed for both TMS and SDMT. The model containing the sex-CAPS interaction provided a superior fit to the data compared to the baseline model. TMS showed significant effects due to sex alone (p < 0.0001) and the sex-CAPS interaction (p < 0.0001), with females having lower TMS scores at low CAPS but a greater CAPS-mediated increase (**Figure 1B**). Significant sex effects and sex-CAPS interactions were also found in SDMT (sex: p < 0.0001, sex-CAPS interaction: p < 0.0001). Similarly, females exhibited better performance at low CAPS and a more pronounced decline with increasing CAPS. Subgroup analyses revealed the sex-CAPS interaction in manifest subjects only (TMS: sex p < 0.05, sex-CAPS p < 0.001; SDMT: sex p < 0.01, sex-CAPS p < 0.01), although significant sex differences remained for SDMT in the premanifest cohort (p < 0.0001).

### HD-ISS Stage 3 landmarks – TFC and Independence Scale

Significant sex-CAPS interactions were found for TFC and Independence Scale (TFC: p < 0.05; Independence Scale: p < 0.001), with females showing a greater CAPS-mediated decline in function (**Figure 1C**). In manifest HD patients, both TFC and Independence Scale were significantly associated with the sex-CAPS interaction (TFC: p < 0.01; Independence Scale: p < 0.0001), and Independence Scale also showed an independent sex effect (p < 0.01), indicating higher function in females when controlling for other factors. In premanifest subjects, the opposite sex effect and sex-CAPS interaction were observed for Independence Scale (sex: p < 0.05; sex-CAPS: p < 0.01).

### Impact of sex on HD-ISS staging

Analysis of HD-ISS Stage assignment showed a general trend of more males being assigned to Stage 2 compared to females with the same CAG repeat length and age range (**Figure 2A**). Chi-square tests revealed significantly higher percentage of males assigned to Stage 2 at several CAG repeat lengths, with up to a 20% difference (**Figure 2B**). Stage 3 assignment (**Supplementary Figure 2**) showed no significant differences between males and females after BH correction.

**Figure 2.**
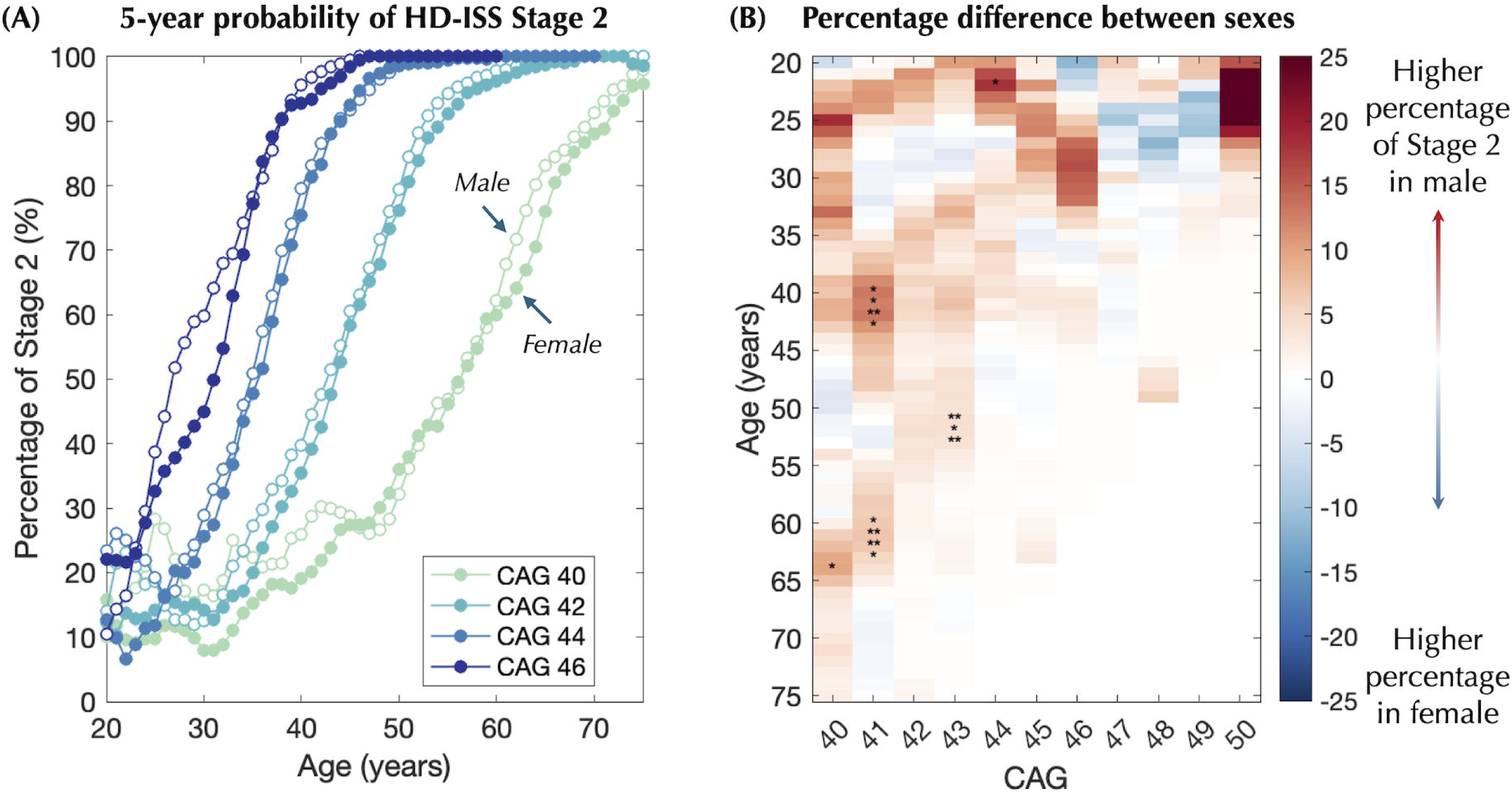
Probability of Stage 2 assignment within a 5-year range for specific CAG repeat lengths (A) are plotted for male and female subjects. The percentage differences are illustrated in (B). Significantly differences in percentage are highlighted with stars.

### Cortical thickness evaluation

Females exhibited significantly greater cortical thickness in the left hemisphere’s supramarginal and postcentral regions, and the right inferior parietal and postcentral regions (**Figure 3A**). The correlation with CAPS indicated significant disease burden-mediated cortical thinning, which was widespread throughout the cerebrum. The sex-CAPS interaction showed region-specific patterns, with a negative interaction (more prominent thinning with increasing disease burden in females) observed in the left hemisphere’s supramarginal gyrus and the left entorhinal cortex. A more widespread positive interaction (less prominent thinning with increasing disease burden in females) was observed in the left superior parietal cortex, right precentral gyrus, and subregions of the inferior and middle temporal gyri and cingulate cortex (**Figure 3B-C**).

**Figure 3.**
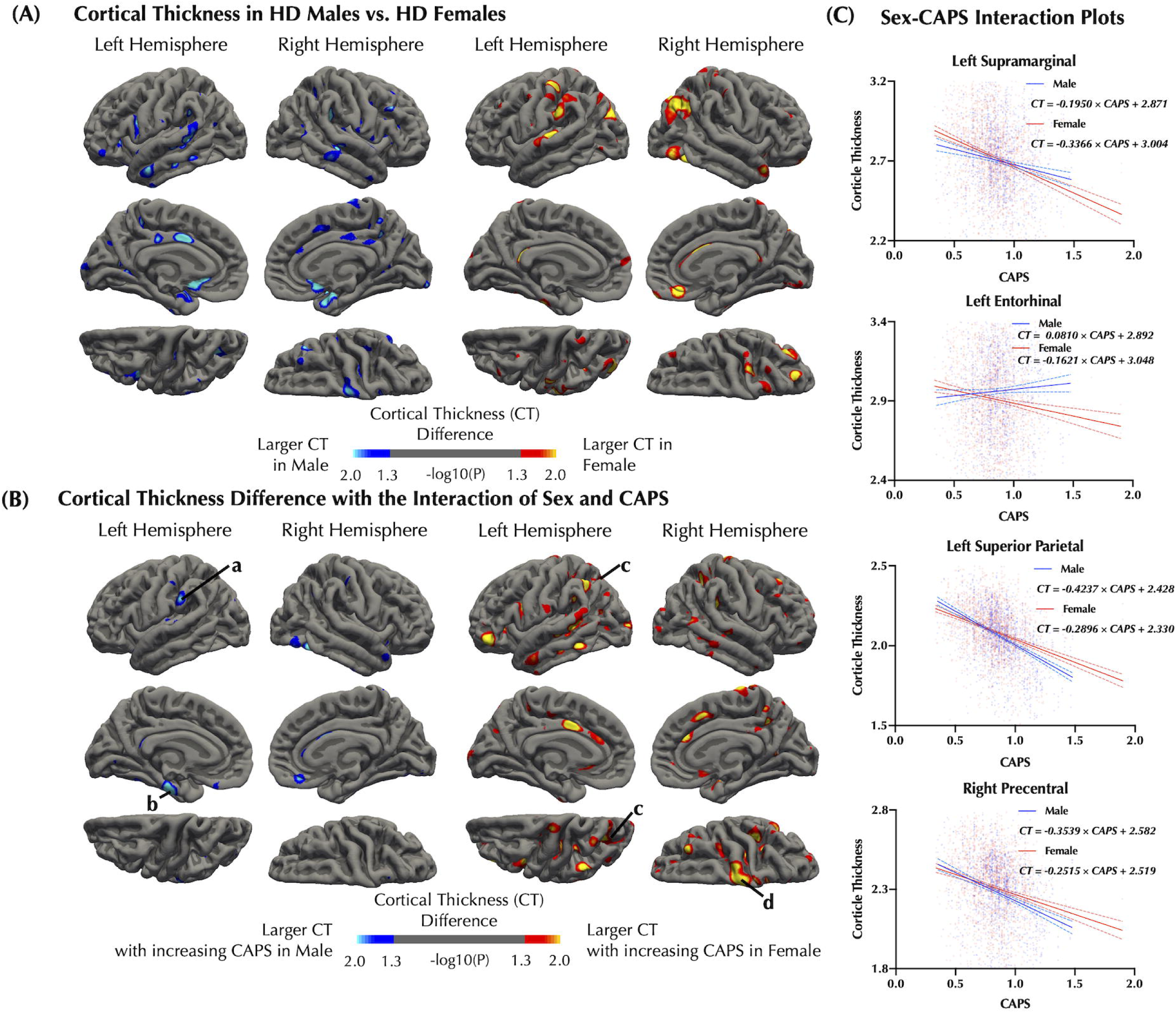
Cortical thickness differences between male and female participants (A) and regarding the sex-CAPS interaction (B) are illustrated. Four clusters of significant cortical regions in sex-CAPS interaction (labeled a-d) are further plotted to demonstrate the sex-CAPS interaction (C). CT: cortical thickness.

### Other neuropsychiatric assessment

No significant sex-CAPS interaction effects were observed on PBA-S scores. Only the depression score was significantly influenced by sex (p < 0.0001), with females exhibiting higher scores. In premanifest subjects, significant effects of both sex and sex-CAPS interaction were observed for depression (sex: p < 0.001, sex-CAPS: p < 0.05) and irritability (sex: p < 0.05, sex-CAPS: p < 0.05). Higher scores were observed in female subjects, with a less degree of CAPS-mediated increase compared to males. In manifest HD subjects, only depression score showed significant dependencies on sex and sex-CAPS interaction (sex: p < 0.0001, sex-CAPS: p < 0.01). Both males and females exhibited a decrease in depression score with increasing CAPS, with females showing a greater degree of CAPS- mediated decrease. Detailed statistics are included in **Table 2** and **Supplementary Tables 3 and 4**.

**Table 2.**
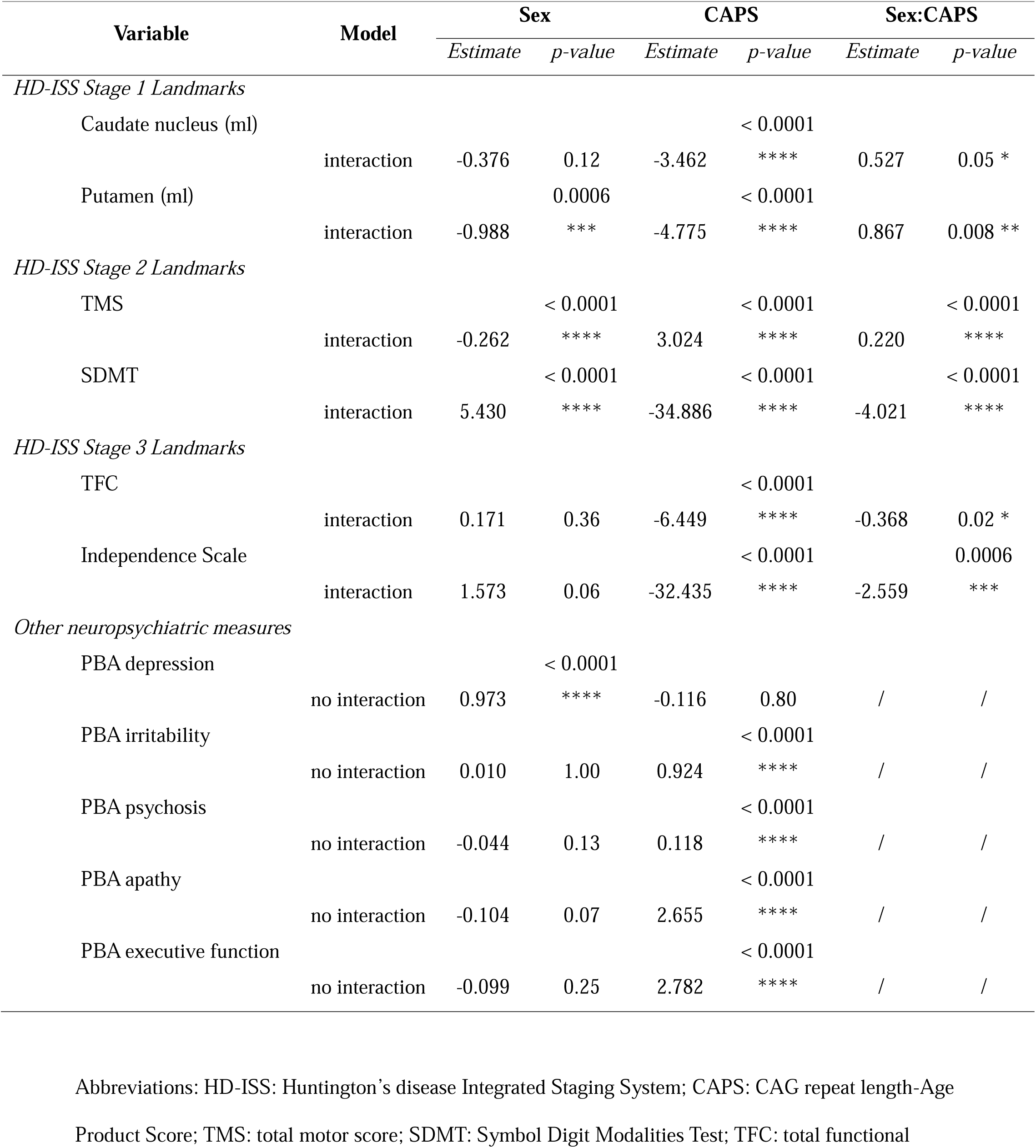
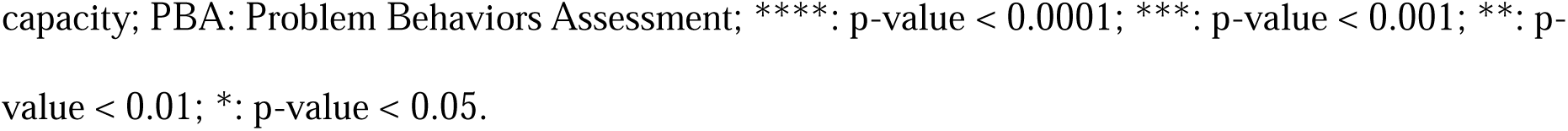
Linear mixed model fitting of clinical variables and brain volumes in all HD subjects.

| Variable | Model | Sex |  | CAPS |  | Sex:CAPS |  |
| --- | --- | --- | --- | --- | --- | --- | --- |
|  |  | Estimate | p-value | Estimate | p-value | Estimate | p-value |
| HD-ISS Stage 1 Landmarks |  |  |  |  |  |  |  |
| Caudate nucleus (ml) |  |  |  |  | < 0.0001 |  |  |
|  | interaction | -0.376 | 0.12 | -3.462 | **** | 0.527 | 0.05 * |
| Putamen (ml) |  |  | 0.0006 |  | < 0.0001 |  |  |
|  | interaction | -0.988 | *** | -4.775 | **** | 0.867 | 0.008 ** |
| HD-ISS Stage 2 Landmarks |  |  |  |  |  |  |  |
| TMS |  |  | < 0.0001 |  | < 0.0001 |  | < 0.0001 |
|  | interaction | -0.262 | **** | 3.024 | **** | 0.220 | **** |
| SDMT |  |  | < 0.0001 |  | < 0.0001 |  | < 0.0001 |
|  | interaction | 5.430 | **** | -34.886 | **** | -4.021 | **** |
| HD-ISS Stage 3 Landmarks |  |  |  |  |  |  |  |
| TFC |  |  |  |  | < 0.0001 |  |  |
|  | interaction | 0.171 | 0.36 | -6.449 | **** | -0.368 | 0.02 * |
| Independence Scale |  |  |  |  | < 0.0001 |  | 0.0006 |
|  | interaction | 1.573 | 0.06 | -32.435 | **** | -2.559 | *** |
| Other neuropsychiatric measures |  |  |  |  |  |  |  |
| PBA depression |  |  | < 0.0001 |  |  |  |  |
|  | no interaction | 0.973 | **** | -0.116 | 0.80 | / | / |
| PBA irritability |  |  |  |  | < 0.0001 |  |  |
|  | no interaction | 0.010 | 1.00 | 0.924 | **** | / | / |
| PBA psychosis |  |  |  |  | < 0.0001 |  |  |
|  | no interaction | -0.044 | 0.13 | 0.118 | **** | / | / |
| PBA apathy |  |  |  |  | < 0.0001 |  |  |
|  | no interaction | -0.104 | 0.07 | 2.655 | **** | / | / |
| PBA executive function |  |  |  |  | < 0.0001 |  |  |
|  | no interaction | -0.099 | 0.25 | 2.782 | **** | / | / |

Abbreviations: HD-ISS: Huntington's disease Integrated Staging System; CAPS: CAG repeat length-Age
Product Score; TMS: total motor score; SDMT: Symbol Digit Modalities Test; TFC: total functional
capacity; PBA: Problem Behaviors Assessment; \*\*\*\*: p-value < 0.0001; \*\*\*: p-value < 0.001; \*\*: p-value < 0.01; \*: p-value < 0.05.

### Longitudinal progression

Sex differences in longitudinal progression were observed in several clinical and MRI-based assessments, as indicated by the interaction between days from baseline and sex. Significant sex-time interactions were found in PBA-S depression score (p < 0.01), SDMT (p < 0.05), and caudate volume (p < 0.05) in all HD subjects (**Supplementary Table 5**). Depression scores and caudate volume atrophy progressed slower in female subjects compared to male subjects, whereas cognitive decline as measured by SDMT was faster in females.

## DISCUSSION

This study investigated the interplay between sex and disease burden on clinical measures and neuroimaging biomarkers using a comprehensive dataset from four HD studies. Our study revealed significant interaction between sex and disease burden, with female HD gene carriers showing more pronounced motor, cognitive, and functional declines with increasing disease burden, yet presenting lower levels of CAPS-mediated striatal atrophy and cortical thinning compared to males. These findings underscore the importance of sex-specific considerations in the clinical management and staging of HD.

Our findings highlight significant sex-specific differences in HD symptomatology, aligning with and expanding upon previous research (**Supplementary Table 5**). Female HD subjects had lower TMS and better SDMT performance at lower disease burden, yet experienced more pronounced increases in TMS and declines in SDMT as disease burden increased. Functional capacity, assessed by TFC and Independence Scale, declined more rapidly in females with increasing CAPS. We hypothesize that the greater symptom severity in female manifest HD subjects may result from a greater susceptibility to disease burden, supported by the superior fit of the sex-CAPS interaction model (Model 2).

We also observed higher depression levels in female HD subjects, consistent with prior reports ^5,6,26,27^. In pre-motor HD patients, depression scores increased with CAPS, although less so in females. In manifest HD patients, depression severity decreased with higher disease burden, corroborating previous studies ^6,26,27^. The non-monotonic CAPS dependence at different disease stages is consistent with reports that depressive symptoms peak during intermediate stage (defined based on TFC) and diminish in later stages ^28^, suggesting the occurrence of anosognosia in females at later disease stages when more severe cognitive impairment also ensues. This also implies that the observed sex difference in depression symptoms in HD is not merely a direct effect of sex but is also significantly influenced by the sex-disease burden interaction.

Our study revealed significant sex-specific differences in neuroimaging biomarkers among HD patients. We found similar caudate volume and smaller putamen volume in female HD gene carriers compared to males after adjusting for intracranial volumes at lower disease burden, aligning with existing literature in healthy individuals ^29–32^. Despite these smaller initial volumes, females showed less pronounced atrophy in these regions with increasing disease burden. Cortical thickness evaluations indicated that females experienced less cortical thinning with increasing disease burden than males. Reports on sex differences in brain atrophy in HD are scarce, despite the use of caudate and putamen measures as landmarks in the HD-ISS system. Our findings fill this knowledge gap, supporting the consideration of sex in HD disease monitoring and clinical trials.

One unexpected finding was the divergence between trends in clinical measures and neuroimaging biomarkers. While clinical measures indicate worse deterioration in females with increasing disease burden, neuroimaging measures suggest relative resilience to structural brain changes in females. This paradox may be explained by neuroprotective factors delaying structural degeneration but not clinical manifestations. Another explanation could be the differences in population characteristics between the entire cohort and the imaging sub-cohort, which had fewer manifest HD patients, potentially biasing imaging findings towards earlier stages of the disease.

The mechanism behind sex differences in sensitivity to disease burden remains unclear. While differential vulnerability to neurodegeneration in males and females is documented in other neurodegenerative diseases ^33^, little is known about biological factors, such as hormonal differences, that influence HD progression. Both estrogen and testosterone have been associated with neuroprotection. In male HD gene carriers, decreased plasma testosterone levels correlate with increased disease severity and cognitive impairment, whereas in female subjects, lower testosterone levels are found in patients with depression ^34,35^. Estrogen’s neuroprotection effect, demonstrated in other neurological diseases ^36^, may explain the less pronounced brain structural degeneration and better motor and cognitive performance at earlier disease stages in female HD subjects, which may decline as the effect of mutant HTT accumulates or during the transition into menopause. Preclinical studies suggest that estrogen may protect against motor impairment and atrophy of medium-sized spiny neurons in transgenic HD rats ^37^, and depressive symptoms in R6/1 HD mice ^38^. However, estrogen treatment has shown limited efficacy in human HD subjects ^39^, likely because other factors, such as sex-specific immune responses ^40^ and sex differences in genotypes and allele frequencies ^41,42^, may also play a role. Further research is needed to elucidate these mechanisms and their implications for clinical management of HD patients.

This study has several limitations that should be acknowledged. The combined dataset from multiple HD studies, while comprehensive, may have inherent inconsistencies in data collection, participant characteristics, and clinical assessments, affecting the uniformity of results. CAPS as a proxy for disease burden, although widely used, may not fully capture disease progression complexity. The imbalanced race/ethnicity distribution, with most samples from Western countries, may limit the generalizability of our findings. Our imaging data analysis is limited to a small subset of the entire cohort (< 10%), which may not capture the full dynamic range in the larger cohort. Lack of manual quality assurance in brain structure parcellation may induce variability. Lastly, our linear mixed model may not fully capture the complexity of symptom and biomarker progression, such as the non-monotonic changes in depressive symptoms. Future studies should aim for greater diversity in participant demographics, validation of imaging findings, and more sophisticated statistical models to fully understand the nuances of sex differences in HD progression.

The observed sex differences in sensitivity to disease burden suggest that sex-specific considerations should be integrated into the HD staging and clinical management. Females may perform better on clinical measures at lower disease burdens, potentially delaying their assignment to more advanced stages. Our analysis indicates a slightly higher but nonetheless significant percentage of male subjects assigned to Stage 2 compared to female subjects within the same age range and CAG repeat length. Despite potential delays in Stage assignment for females, their faster progression at higher disease burdens highlights the need for earlier intervention and closer monitoring. Investigational disease-modifying treatments may also require different cut-offs based on different sex and disease stages, given their interaction on clinical and imaging biomarkers. These insights emphasize the importance of incorporating sex as a critical factor in HD research, clinical trials, and patient management protocols to enhance the overall patient care.

## ACKNOWLEDGEMENT

Data used in this work were generously provided by the participants in the Enroll-HD, PREDICT-HD, TRACK-HD/ON, and IMAGE-HD studies and made available by CHDI Foundation, Inc. Enroll-HD is a clinical research platform and longitudinal observational study for Huntington’s disease families intended to accelerate progress towards therapeutics; it is sponsored by CHDI Foundation, a nonprofit biomedical research organization exclusively dedicated to collaboratively developing therapeutics for HD. Enroll-HD would not be possible without the vital contribution of the research participants and their families.

## AUTHORS’ ROLES

Research Project: A. Conception, B. Organization, C. Execution; 2. Statistical Analysis: A. Design, B. Execution, C. Review and critique; 3. Manuscript Preparation: A. Writing of the first draft, B. Review and critique.

J.Y.: 1A, 1B, 1C, 2A, 2B, 3A

G.F.: 1C, 2B, 3A

G.S.: 1C, 2B, 3A

A.M.: 1A, 2C, 3B

D.T.: 2A, 2C, 3B

K.F.: 1B, 2B

L.I.: 1B, 2B

J.S.: 1B, 2B

M.A.M: 1C, 2C, 3B

J.M.L.: 2C, 3B

## FINANCIAL DISCLOSURES OF ALL AUTHORS (FOR THE PRECEDING 12 MONTHS)

A.M. has provided consultation services to Neurocrine Biosciences in 2023. A.M. receives funding from the Huntington Disease Society of America (HDSA) and Berman Topper Career Development Fellowship.

