## Supplementary for "Interplay between Sex and Disease Burden in Huntington’s Disease: Clinical and Neuroimaging Perspectives"

Supplementary Table 1. HD dataset information

|  | **ENROLL-HD** | **PREDICT-HD** | **TRACK-HD/ON** | **IMAGE-HD** |
| --- | --- | --- | --- | --- |
| Study Objectives | Observational cohort study to identify factors affecting HD progression. | Observational study to identify biological and clinical markers of pre-HD; validate the optimal markers for preventive clinical trials. | Observational study to examine clinical and biological findings of disease progression. | Observational longitudinal multi-modal neuroimaging study to identify brain changes in HD. |
| Geographical Coverage | >20 countries across North America, Latin America, Europe, and Australasia | 33 sites across the US, Canada, UK, Germany, Spain, and Australia | 4 sites in UK, France, Netherlands, and Canada | Single site in Australia |
| Participant Criteria | HD gene carriers, family members, genotype-negative individuals | HD gene carriers (case), controls | Early-stage HD patients, premanifest HD gene carriers, controls | Premanifest and early-stage HD, controls |
| Data Collection Frequency | Annual visits | Annual visits (up to 14 visits) | Annual visits (up to 7 visits) | Baseline, 18 months, 30 months |
| Core Data Elements | Demographics, UHDRS, cognitive, motor, behavioral, functional assessments, CAG-repeat length | Demographics, UHDRS, Neuropsychological assessment, MRI, CAG-repeat length | Demographics, UHDRS, cognitive, motor, behavioral, functional assessments, MRI, CAG-repeat length | Demographics, UHDRS MRI, cognitive assessments, MRI, CAG-repeat length |
| Premanifest vs Manifest HD | Based on clinical assessment | Only premanifest HD is included | Premanifest HD: TMS <= 5 and DCL < 4 | Premanifest HD: TMS <= 5 |
| Sample Size | >25,550 participants | 1,485 participants | 446 participants | 108 participants |
| Race/Ethnicity Groups | Caucasian, American Black, Hispanic or Latino Origin, American Indian / Native American / Amerindian, Asian, Mixed, Other | White, Black or African American, American Indian / Alaska Native, Asian, More than one race, Unknown; Hispanic / Latino, Not Hispanic / Latino | Caucasian, African – Black, African – North, Asian – West, Asian – East, American – Black, American – Latin, Mixed, Other, Unknown | N/A |
| Status | Ongoing | Completed | Completed | Completed |

Note: N/A indicates data not reported by the studies.

Supplementary Table 2. Representative T_1_-weighted sequence parameters

|  | **IMAGE-HD** | **TRACK-HD/ON** | **PREDICT-HD** |
| --- | --- | --- | --- |
| Sequence | 3D MPRAGE | 3D MPRAGE | 3D SPGR |
| Scanner/Vendor | Siemens Trio | Siemens | N/A |
| Matrix Size | 320 x 320 | 256 x 256 | 256 x 192 |
| Field of View (mm) | 270 x 270 | 280 x 280 | 240 x 180 |
| Number of Slices | 192 | 208 | 124 |
| Slice Thickness (mm) | 0.9 | 1.0 | 1.5 |
| TR (ms) | 1900 | 2200 | 18 |
| TE (ms) | 2.59 | 2.2 | 3 |
| TI (ms) | 900 | N/A | N/A |
| Number of Averages | N/A | N/A | 2 |
| Flip Angle | 9° | 10° | 20° |
| Field Strength | 3T | 3T | 1.5T |

Note: N/A indicates parameters not reported by the studies.

Supplementary Table 3. Linear mixed model fitting of clinical variables and brain volumes in premanifest HD subjects

| Variable | Model | Sex | | CAPS | | Sex:CAPS | |
| --- | --- | --- | --- | --- | --- | --- | --- |
|  |  | *Estimate* | *p-value* | *Estimate* | *p-value* | *Estimate* | *p-value* |
| *HD-ISS Stage 1 Landmarks* |  |  |  |  |  |  |  |
| Caudate nucleus (ml) | no interaction | 0.093 | 0.20 | -2.666 | < 0.0001 **** | / | / |
| Putamen (ml) | no interaction | -0.210 | 0.01 * | -3.780 | < 0.0001 **** | / | / |
| *HD-ISS Stage 2 Landmarks* |  |  |  |  |  |  |  |
| TMS | no interaction | -0.023 | 0.40 | 1.399 | < 0.0001 **** | / | / |
| SDMT | no interaction | 2.406 | < 0.0001 **** | -20.766 | < 0.0001 **** | / | / |
| *HD-ISS Stage 3 Landmarks* |  |  |  |  |  |  |  |
| TFC | no interaction | 0.015 | 0.82 | -0.394 | < 0.0001 **** | / | / |
| Independence Scale | interaction | -0.886 | 0.04 * | -2.111 | < 0.0001 **** | 1.371 | 0.01 ** |
| *Other neuropsychiatric measures* |  |  |  |  |  |  |  |
| PBA depression | interaction | 2.028 | 0.0002 *** | 2.880 | < 0.0001 **** | -1.405 | 0.05 * |
| PBA irritability | interaction | 0.880 | 0.02 * | 1.250 | 0.001 ** | -0.986 | 0.02 * |
| PBA psychosis | no interaction | -0.039 | 0.23 | 0.116 | 0.24 | / | / |
| PBA apathy | no interaction | -0.092 | 0.25 | 0.997 | < 0.0001 **** | / | / |
| PBA executive function | interaction | 0.621 | 0.05 | 1.171 | 0.0007 *** | -0.859 | 0.03 * |

Supplementary Table 4. Linear mixed model fitting of clinical variables and brain volumes in manifest HD subjects

| Variable | Model | Sex | | CAPS | | Sex:CAPS | |
| --- | --- | --- | --- | --- | --- | --- | --- |
|  |  | *Estimate* | *p-value* | *Estimate* | *p-value* | *Estimate* | *p-value* |
| *HD-ISS Stage 1 Landmarks* |  |  |  |  |  |  |  |
| Caudate nucleus (ml) | interaction | -1.461 | 0.03 * | -3.587 | < 0.0001 **** | 1.399 | 0.02 * |
| Putamen (ml) | interaction | -1.752 | 0.04 * | -4.350 | < 0.0001 **** | 1.324 | 0.08 |
| *HD-ISS Stage 2 Landmarks* |  |  |  |  |  |  |  |
| TMS | interaction | -0.115 | 0.01 * | 1.376 | < 0.0001 **** | 0.136 | 0.0005 *** |
| SDMT | interaction | 3.261 | 0.003 ** | -24.200 | < 0.0001 **** | -2.273 | 0.003 ** |
| *HD-ISS Stage 3 Landmarks* |  |  |  |  |  |  |  |
| TFC | interaction | 0.593 | 0.08 | -6.815 | < 0.0001 **** | -0.777 | 0.004 ** |
| Independence Scale | interaction | 4.682 | 0.003 ** | -34.798 | < 0.0001 **** | -5.402 | < 0.0001 **** |
| *Other neuropsychiatric measures* |  |  |  |  |  |  |  |
| PBA depression | interaction | 2.578 | < 0.0001 **** | -2.698 | < 0.0001 **** | -1.233 | 0.007 ** |
| PBA irritability | no interaction | 0.038 | 0.75 | -0.928 | < 0.0001 **** | / | / |
| PBA psychosis | no interaction | -0.031 | 0.45 | -0.167 | 0.06 | / | / |
| PBA apathy | no interaction | -0.065 | 0.43 | 1.533 | < 0.0001 **** | / | / |
| PBA executive function | no interaction | -0.065 | 0.63 | 1.585 | < 0.0001 **** | / | / |

Supplementary Table 5**.** Linear mixed model fitting of clinical variables and brain volumes in all HD subjects

| Variable | Sex | CAPS | Sex:  CAPS | Age  (yr) | Days  (yr) | Days:  CAPS | Days:  Sex |
| --- | --- | --- | --- | --- | --- | --- | --- |
| *HD-ISS Stage 1 Landmarks* |  |  |  |  |  |  |  |
| Caudate nucleus (ml) | -0.376 | -3.462 **** | 0.527 * | -0.002 | -0.090 **** | -0.021 | 0.017 * |
| Putamen (ml) | -0.988 *** | -4.775 **** | 0.876 ** | -0.014 **** | -0.144 **** | -0.009 | 0.008 |
| *HD-ISS Stage 2 Landmarks* |  |  |  |  |  |  |  |
| TMS | -0.262 **** | 3.024 **** | 0.220 **** | 0.020 **** | 0.042 **** | 0.050 **** | 0.001 |
| SDMT | 5.430 **** | -34.886 **** | -4.021 **** | -0.240 **** | 1.851 **** | -2.987 **** | -0.090 * |
| *HD-ISS Stage 3 Landmarks* |  |  |  |  |  |  |  |
| TFC | 0.171 | -6.449 **** | -0.368 * | -0.036 **** | 0.508 **** | -0.912 **** | 0.004 |
| Independence Scale | 1.573 | -32.435 **** | -2.559 *** | -0.154 **** | 3.263 **** | -5.429 **** | -0.064 |
| *Other neuropsychiatric measures* |  |  |  |  |  |  |  |
| PBA depression | 0.973 **** | -0.116 | / | -0.006 | 0.033 | -0.163 ** | -0.074 ** |
| PBA irritability | 0.010 | 0.924 **** | / | -0.004 | -0.014 | 0.011 | -0.033 |
| PBA psychosis | -0.044 | 0.181 **** | / | -0.001 | -0.011 | 0.008 | -0.002 |
| PBA apathy | -0.104 | 2.655 **** | / | 0.019 **** | -0.273 **** | 0.421 **** | -0.014 |
| PBA executive function | -0.099 | 2.782 **** | / | -0.004 | -0.165 *** | 0.292 **** | -0.011 |

Supplementary Table 6**.** Summary of the existing literature on sex differences in HD clinical measures.

| Study | Dataset | Subject population | Main Findings | Statistical Model |
| --- | --- | --- | --- | --- |
| Epping et al. (2013) | PREDICT-HD | Prodromal HD (N = 803); Controls (N = 233) | Increased depressive symptoms were significantly associated with female sex. | Chi-square across BDI symptom level groups |
| Hentosh et al. (2021) | ENROLL-HD | Manifest HD (N = 8401) | Female patients presented more severe motor and cognitive decline and worse depression symptoms. | Linear mixed model with sex and CAPS as covariates |
| Zielonka et al. (2013) | EHDN | Manifest HD (N = 1267) | A faster rate of progression in women in the functional assessment, the motor assessment, and the independence scale. | Linear mixed model with age of onset, disease burden, disease duration, smoking status, alcohol abuse, depression, and the number of years of education as covariates |
| Zielonka et al. (2018) | REGISTRY | Manifest HD (N = 2191) | Motor symptoms correlated more with functional ability and influenced function variability more in women than in men. | Simultaneous linear regression with least-square fit method |
| Rocha et al. (2022) | ENROLL-HD | HD gene carriers (N = 11,582) | Female sex was described as a significant predictor of depression among HD gene expansion carriers. | Binary logistic regression model |

Abbreviations: EHDN: European Huntington's Disease Network.

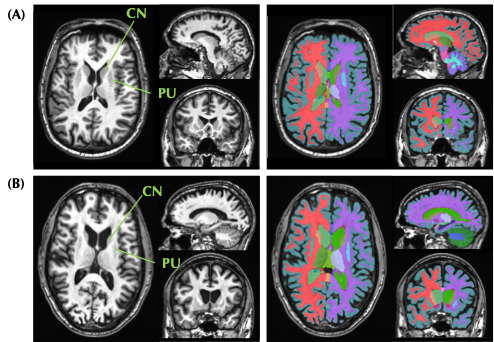

**Supplementary Figure 1.** Example brain region segmentations from one premanifest HD subject (A) and one manifest HD subject (B) in the TRACK-HD dataset. CN: caudate nucleus; PU: putamen.

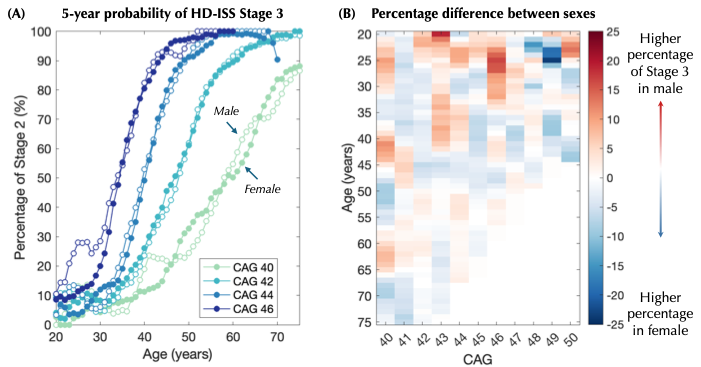

**Supplementary Figure 2.** Percentage of subjects assigned to HD-ISS Stage 3 compared between male and female HD gene carriers. Probability of Stage 2 assignment within a 5-year range for specific CAG repeat lengths (A) are plotted between male and female subjects. The percentage differences are illustrated in (B). No significantly differences in percentage were observed after BH correction.
